# A Comprehensive, Low-Cost Multistation ENT Simulation Curriculum for Medical Students: Five Reproducible Task Trainers for Foundational Otolaryngology Skills

**DOI:** 10.64898/2026.05.18.26353510

**Authors:** Tanner J. Jefferies, Mark K. Lavigne

**Affiliations:** Campbell University School of Osteopathic Medicine, Surgery Department, Lillington, NC, USA; Scotland Memorial Hospital, Department of Otolaryngology, Laurinburg, NC, USA

**Keywords:** Otolaryngology education, Simulation-based education, Undergraduate medical education, Low-cost simulation, Procedural skills training

## Abstract

**Introduction:** Early exposure to otolaryngology (ENT) procedural skills in undergraduate medical education is limited by patient safety concerns, restricted clinical opportunities, and the cost of commercial simulators. As a result, essential ENT skills are often underrepresented in structured, hands-on curricula for medical students.

**Methods:** We developed a low-cost, multistation ENT simulation curriculum consisting of five reproducible task trainers: ear examination and otologic procedures, mirror laryngoscopy, rigid and flexible endoscopic navigation, introductory mastoid drilling, and emergency cricothyrotomy. The curriculum was delivered as a 2-hour, faculty-led workshop during a third-year medical student otolaryngology rotation. Learners rotated through stations in small groups. Pre- and post-workshop surveys assessed self-reported anatomical familiarity, procedural confidence, and educational value using a 5-point Likert scale, with additional qualitative feedback collected.

**Results:** All participants completed pre- and post-workshop evaluations. Learners demonstrated increased confidence across all assessed anatomical and procedural domains, including otoscopy, endoscopy, mirror laryngoscopy, mastoid drilling orientation, and cricothyroid membrane identification. Educational value ratings were high across all stations, with mean scores ranging from 4.33 to 5.00. Qualitative feedback emphasized the realism, accessibility, and benefit of hands-on practice in a low-stakes learning environment.

**Conclusion:** This low-cost, multistation ENT simulation curriculum provides a feasible and reproducible approach for introducing foundational otolaryngology skills to medical students. The structured format and affordable models support early procedural exposure and may enhance learner preparedness prior to supervised clinical encounters, particularly in settings with limited simulation resources.

## Introduction

Otolaryngology-related procedures—such as otoscopy, nasal endoscopy, mirror laryngoscopy, cricothyrotomy, and mastoidectomy drilling—constitute essential skills in primary care, emergency medicine, and surgical practice. However, early learners often face limited opportunities to practice these skills due to patient discomfort, safety concerns, and restricted access to cadaveric materials or high-fidelity commercial simulators, which are frequently cost-prohibitive and challenging to obtain [1-4]. Consequently, medical students typically encounter ENT examinations and procedures for the first time during clinical rotations, where real-time learning is hampered by patient safety considerations and time constraints [1-2].

Simulation-based education provides a safe and controlled environment for developing procedural competence, enhancing psychomotor skills, and improving spatial understanding. Robust evidence illustrates that simulation accelerates skill acquisition, boosts confidence, and reduces errors during clinical performance [5-7]. In the field of otolaryngology, there is a growing interest in accessible, low-cost simulation methods, leading to the development of reproducible training tools for sinus surgery, otomicroscopy, phonomicrosurgery, mastoid drilling, and fundamental surgical skills [8-12]. These innovations have lowered financial and logistical barriers, facilitating repeated, deliberate practice—an essential component in achieving procedural mastery [11,12].

Despite the presence of several low-cost models, there is a notable lack of comprehensive curricula that effectively integrate multiple ENT task trainers into a cohesive educational framework. Most existing models are presented as isolated technical reports rather than as components of a structured instructional sequence. To bridge this gap, we developed a multi-station, low-cost ENT simulation curriculum that incorporates five reproducible task trainers discussed in the literature: an ear model for otoscopy and tympanic membrane procedures [9], a mirror laryngoscopy trainer with pathology stations [12], a nasal and laryngeal endoscopy model [3], a cortical mastoidectomy drilling trainer [9], and an ultra-low-cost cricothyrotomy model that emphasizes landmark recognition and airway access [7].

This curriculum was implemented as a two-hour ENT simulation workshop for medical students, aiming to introduce essential procedural skills prior to supervised clinical encounters. The purpose of this report is to describe the curriculum structure, present learner outcomes through pre/post evaluation measures, and provide fully reproducible materials to facilitate the integration of low-cost ENT simulation into undergraduate medical education.

## Methods

### Curriculum Development

This curriculum was designed to familiarize preclinical and clinical medical students with fundamental otolaryngology procedures through a series of low-cost and reproducible task trainers developed by the authors. The curriculum features five distinct models: (1) a trainer for ear examination and tympanic membrane repair, (2) a mirror laryngoscopy trainer with interchangeable pathology stations, (3) a nasal and laryngeal endoscopy trainer, (4) a cortical mastoidectomy drilling trainer, and (5) an ultra-low-cost cricothyrotomy trainer.

Of these models, only the cortical mastoidectomy trainer has previously been documented in a technical report that outlines its design and educational effectiveness [9]. The other models were newly developed by the authors to address specific skill gaps often encountered in early ENT education. Each model was purposely crafted to be cost-effective, easily assembled, and suitable for deliberate practice, consistent with research that emphasizes the importance of low-cost simulation in early procedural training. The curriculum is structured as a two-hour, multi-station workshop to optimize hands-on experience and faculty-guided coaching. To support reproducibility and dissemination, detailed build guides for each task trainer, facilitator materials, learner task sheets, and evaluation instruments are provided in the appendices (Appendices A–I).

### Participants

The participants in this study were third-year medical students who were undergoing their otolaryngology rotation at Scotland Memorial Hospital. The simulation workshop was incorporated into the structured educational curriculum that is regularly provided during this rotation. All students assigned to the service were required to take part in the session as a fundamental part of their clinical education.

During the workshop, learners engaged in small groups of three to four students and rotated through five different simulation stations, all guided by otolaryngology faculty members. As part of the standard program evaluation procedures aimed at enhancing educational quality at the rotation site, anonymous pre- and post-session evaluations were collected. No identifying information was gathered.

### Equipment and Materials

All simulation models were constructed using affordable and readily available materials to ensure both accessibility and reproducibility. The cost of materials varied, with prices starting at approximately $1.30 for the mirror laryngoscopy trainer and reaching up to $27.00 for the mastoidectomy model (Appendix I). Standard otolaryngology instruments were utilized at each station, including cerumen curettes, alligator forceps, 0° rigid nasal endoscopes, flexible nasolaryngoscopes, laryngeal mirrors, and a handheld rotary drill for the mastoidectomy station. Comprehensive step-by-step construction guides for each simulation model are included in Appendices A– E.

This combination of stations ensured that learners practiced a balanced set of skills, including fine-motor control, three-dimensional orientation, instrumentation technique, and emergency airway access.

### Curriculum Structure and Implementation

#### Workshop Format

The simulation curriculum was delivered as a two-hour, multistation workshop integrated into the otolaryngology rotation’s scheduled educational didactics. The session followed a structured sequence consisting of:

1. **Orientation and Introduction (10 minutes)**
  - Overview of the goals of the workshop
  - Demonstration of basic ENT instruments
  - Explanation of each simulation station
  - Pre-workshop survey
2. **Five Rotating Simulation Stations (75–90 minutes)**
  - Small groups rotated every 15–20 minutes
  - Faculty provided demonstration, coaching, and real-time feedback
3. **Group Debrief and Discussion (10 minutes)**
  - Reinforcement of key concepts
  - Opportunity to ask questions
  - Reflection on procedural confidence and challenges
  - Post-workshop survey

The debriefing was conducted in a facilitated, group-based format that emphasized key anatomical concepts, procedural steps, and reflection from the learners. This approach aligns with best practices in simulation-based education.

Appendix G includes a detailed facilitator guide with specific teaching points for each station, safety considerations, and troubleshooting strategies. Learner-facing task sheets and self-assessment checklists for each station are provided in Appendix H.

### Personnel and Setting

The workshop was led by faculty members from the otolaryngology department, with one instructor assigned to each simulation station. Sessions took place in a skills laboratory or a designated teaching space, outfitted with basic tabletop workstations and standard otolaryngology instruments. The absence of high-fidelity mannequins, dedicated simulation staff, or specialized simulation facilities enabled the curriculum to be executed in regular educational settings.

### Station Procedures

## Station 1: Ear Procedures ures

### Learners practiced

□ Otoscopic visualization of simulated tympanic membranes
□ Removal of mock cerumen
□ Foreign body extraction
□ Tympanic membrane patching or ventilation tube placement

This station emphasized depth perception, gentle instrument handling, and anatomical landmark recognition.

## Station 2: Mirror Laryngoscopy

### Learners practiced

□ Warming and positioning the laryngeal mirror
□ Depressing the simulated tongue
□ Aligning a light source
□ Visualizing printed normal and pathological laryngeal images

Interchangeable laryngeal pathology stations allowed varied diagnostic practice.

## Station 3: Nasal and Laryngeal Endoscopy

### Learners used both rigid and flexible scopes to

□ Navigate the nasal cavity
□ Advance into a simulated nasopharynx
□ Visualize the “larynx” via tubing
□ Remove small simulated foreign bodies

The bell pepper model provided realistic anatomical angles and variable internal anatomy.

## Station 4: Mastoid Drilling

### Using the previously published cortical mastoidectomy model [9], learners practiced

□ Holding and stabilizing a handheld drill
□ Drilling in smooth sweeping motions
□ Identifying the boundaries of McEwen’s triangle
□ Avoiding embedded representations of critical structures

This station introduced drilling mechanics, spatial orientation, and early otologic surgery principles.

## Station 5: Cricothyrotomy

### Learners practiced

□ Palpating the thyroid cartilage, cricothyroid membrane, and tracheal rings
□ Making a horizontal incision through layered simulated tissues
□ Entering a simulated airway
□ Inserting a tube or catheter into the lumen

This station emphasized landmark recognition, tactile feedback, and the sequence of emergency airway access. See Figure 1 for a flow chart of the simulation curriculum.

### Assessment and Evaluation

Anonymous pre- and post-workshop surveys were administered as part of the standard educational program evaluation at Scotland Memorial Hospital. Surveys assessed:

□ Familiarity with ENT anatomy
□ Confidence performing core ENT procedures
□ Confidence in identifying key landmarks
□ Comfort with ENT instruments
□ Perceived educational value of each station

All questions were scored on a 5-point Likert scale. Several open-ended items elicited qualitative feedback on station usefulness, realism of models, and suggestions for improvement.

Survey content was informed by established principles of simulation-based education and previous research on low-cost simulation for procedural training. Pre- and post-workshop responses were analyzed descriptively to evaluate changes in confidence and perceived competence. The complete pre- and post-workshop evaluation instruments are provided in Appendix F.

### Ethical Considerations

This educational activity was reviewed and determined to constitute program evaluation and non–human subjects research. Participation in surveys was voluntary and anonymous, and no identifiable data were collected. The project was conducted in accordance with institutional guidelines and the principles of the Declaration of Helsinki.

## Results

A total of three third-year medical students participated in the workshop as part of their scheduled ENT rotation didactics. All participants completed both the pre- and post-workshop surveys.

### Quantitative Outcomes

Participants in the simulation workshop exhibited significant enhancements in their self-reported familiarity with otolaryngologic anatomy and their confidence in executing fundamental ENT procedures (refer to Tables 1 and 2). Improvements were apparent across all assessed anatomical regions, including the tympanic membrane, nasal cavity, laryngeal structures, anterior neck landmarks, and temporal bone orientation. The most considerable gains in anatomical familiarity were noted particularly in the orientation of the temporal bone and mastoid (see Table 1).

Moreover, confidence in procedural skills improved across all areas, with particularly notable advancements in mirror laryngoscopy, mastoid drilling orientation, the steps involved in cricothyrotomy, and both rigid and flexible endoscopic techniques (see Table 2). These findings indicate that the structured, hands-on simulation curriculum effectively facilitated rapid skill acquisition and significantly enhanced learners’ confidence in diagnostic, endoscopic, surgical, and emergency airway procedures.

### Educational Value of Each Station

#### Learners rated all five stations highly, with mean scores ranging from 4.33 to 5.00 on a 5-point scale

□ Ear Simulation Model: 4.33
□ Nasal & Laryngeal Endoscopy Model: 4.67 D Mirror Laryngoscopy Model: 5.00
□ Cricothyrotomy Model: 5.00
□ Mastoidectomy Model: 5.00

These ratings suggest that each component of the curriculum contributed meaningful educational value.

### Qualitative Feedback

Learner feedback was consistently positive, highlighting the curriculum’s realism, accessibility, and hands-on approach (Table 3). Participants underscored the perceived value of practical experience, with one individual describing the mastoidectomy station as “the most helpful,” emphasizing the importance of skill acquisition through drilling practice. Additionally, learners appreciated the variety of models and stations, noting that exposure to different hands-on activities allowed them to develop a wide range of otolaryngology skills that are typically not encountered early in clinical training.

Participants praised the construction of the models, characterizing the simulators as “realistic,” “approachable,” and “excellent for grasping the fundamentals.” In particular, the bell pepper endoscopy station was recognized for enhancing spatial orientation and foundational endoscopic skills. Suggestions for improvement were minor and focused mainly on technical adjustments, such as incorporating a more rigid internal structure for the pipe cleaners used in the mastoid model and offering alternative otologic instruments to aid the practice of fine motor control.

Overall, students conveyed a strong enthusiasm for the curriculum, indicating that the workshop significantly boosted their confidence ahead of supervised patient interactions.

## Discussion

This multistation ENT simulation curriculum offered third-year medical students early exposure to vital otolaryngology skills through a series of cost-effective and easily reproducible task trainers developed by the authors. Learners exhibited significant increases in self-reported confidence and anatomical familiarity across all assessed domains (refer to Tables 1 and 2). These findings are consistent with previous research indicating that simulation-based education accelerates skill acquisition, enhances learner comfort, and improves procedural readiness [5–7]. Notable improvements were observed across diagnostic, endoscopic, surgical, and emergency airway skills, highlighting the effectiveness of a structured, hands-on approach for novice learners. The combination of quantitative improvements and consistently high ratings of educational value suggests that even brief, small-group simulation experiences can deliver substantial educational benefits early in training.

### Integration of Low-Cost Models Across ENT Domains

An expanding body of literature has detailed individual low-cost ENT simulators, including trainers for sinus surgery, models for phonomicrosurgery, simplified otomicroscopy simulators, and renewable endoscopic skills trainers designed to facilitate repeated practice [8-10,13]. While these models have enhanced access to procedural training, they are frequently presented as standalone technical innovations rather than as integral components of a comprehensive educational curriculum.

In contrast, this curriculum integrates five low-cost, reproducible task trainers into a cohesive multistation experience that covers essential otolaryngology domains, including otologic examination, mirror laryngoscopy, both rigid and flexible endoscopic navigation, principles of mastoid drilling, and emergency front-of-neck access.

Among the included models is the cortical mastoidectomy drilling trainer, which has been previously published by the authors and demonstrated both feasibility and educational utility for early otologic training [9]. Building on this previous work and the broader literature surrounding low-cost simulation, the current study advances the field by organizing multiple task trainers into a structured curriculum that aligns with contemporary recommendations for ENT education [15,16]. This integrated approach directly addresses calls for cohesive, competency-oriented curricula that progressively introduce foundational principles and techniques for novice learners.

### Educational Impact

The workshop’s success in promoting rapid and sustained increases in learner confidence underscores the importance of hands-on ENT simulation in addressing prevalent gaps in undergraduate medical education. Significant improvements were noted across all assessed procedural skills, including mirror laryngoscopy, both rigid and flexible endoscopy, mastoid drilling orientation, and emergency airway procedures (see Table 2). These results align with previous research indicating that structured, low-cost simulation curricula can enhance not only learner confidence but also the effective transfer of procedural skills to clinical and operative practice [14]. Considering that medical students typically have limited exposure to ENT instrumentation before their clinical rotations, early simulation-based training may enhance both engagement and preparedness for supervised clinical involvement.

Crucially, the inclusion of emergency cricothyrotomy training corresponds with the guidelines set forth by the Difficult Airway Society, which stress the early identification of airway failure and familiarity with front-of-neck access techniques as essential aspects of airway management [17]. By integrating airway training into an introductory ENT curriculum, we reinforce patient safety principles while providing students with the opportunity to practice landmark identification and procedural sequencing in a controlled, low-stakes environment.

### Qualitative Feedback and Learner Experience

The qualitative feedback offered valuable insights that complement the quantitative results, with learners consistently praising the realism, accessibility, and educational value of hands-on practice across various stations (refer to Table 3). Participants particularly appreciated the chance to perform procedures they had previously only observed, such as mastoid drilling and flexible nasolaryngoscopy. Many learners reported enhancements in spatial awareness and instrument manipulation, indicating that well-designed, low-fidelity models can effectively aid in the development of foundational psychomotor and cognitive skills.

Suggestions for improvement were minimal and primarily focused on technical enhancements, such as reinforcing certain model components or increasing the range of available instruments. Overall, the positive qualitative feedback highlights the acceptability of the curriculum and reinforces its effectiveness as an introductory platform for ENT procedural education.

### Feasibility, Scalability, and Reproducibility

A key strength of this curriculum lies in its feasibility and affordability. All task trainers have been constructed using low-cost, readily available materials, with expenses ranging from approximately $1.30 to $27.00 per model (Appendix I). This minimal financial barrier promotes scalability and facilitates adoption in institutions that may have limited access to simulation centers or high-fidelity commercial models. The modular design enables the curriculum to be implemented in full or tailored to specific educational contexts, such as clerkship didactics, skills workshops, or procedural boot camps.

The reproducibility of the models further enhances their educational value. Since the trainers can be easily assembled and disassembled, learners can engage in repeated practice over time, which is a crucial aspect of deliberate practice and motor skill acquisition. This repetition allows students to refine their techniques, develop muscle memory, and internalize procedural steps without the pressures associated with patient care, thereby extending the learning experience beyond a single instructional session.

### Limitations

Several limitations warrant careful consideration. The outcomes largely relied on self-reported measures of confidence and familiarity rather than on objective performance evaluations, and the small sample size restricts generalizability. Moreover, the simplified anatomical representations of the models do not capture the full complexity of cadaveric specimens or high-fidelity simulators. Therefore, this curriculum is not designed to replace advanced simulation modalities or comprehensive procedural training.

Instead, it should be regarded as an introductory stepping stone for early learners. By offering structured exposure to instrumentation, procedural flow, and foundational techniques, the curriculum equips students to engage more effectively with high-fidelity simulators and supervised clinical experiences later in their training. Within this specific context, the curriculum fulfills its intended purpose as a foundational educational framework.

### Future Directions

Future research should incorporate objective performance metrics, such as structured checklists or global rating scales, to rigorously evaluate skill acquisition and technical proficiency. Expanding to larger cohorts and implementing multi-institutional studies would enhance the generalizability of findings and facilitate comparative analyses of educational outcomes. Longitudinal assessments may further elucidate skill retention and its transfer to clinical performance.

Additionally, future directions should include the expansion of the curriculum to integrate advanced ENT simulations or hybrid approaches that combine low-cost task trainers with higher-fidelity or virtual simulation platforms as learners advance. This graduated strategy aligns with competency-based medical education and fosters a continuum of procedural training from novice to advanced learner.

## Conclusion

This study outlines the development and implementation of a comprehensive, cost-effective, multistation ENT simulation curriculum aimed at introducing foundational otolaryngology skills to medical students. Through five reproducible task trainers, the curriculum offered structured, hands-on experience in diagnostic, endoscopic, surgical, and emergency airway procedures that are often underrepresented in undergraduate medical education. Participation in the workshop correlated with increased learner confidence, enhanced anatomical familiarity, and a high perceived educational value across all stations, reinforcing the effectiveness of simulation as a valuable adjunct to early procedural training.

By emphasizing affordability, accessibility, and ease of replication, this curriculum presents a practical and scalable method for incorporating ENT procedural education into clerkships or skills-based workshops, particularly in environments with limited simulation resources. Although the results were primarily derived from self-reported measures from a small cohort, the findings indicate that even brief, structured simulation experiences can enhance learners’ preparedness for supervised clinical encounters. Future research should include objective performance assessments, larger sample sizes, and longitudinal follow-up to further assess educational impact and skill retention.

## Data Availability

All data produced in the present study are available upon reasonable request to the authors

## Statements and Declarations

On behalf of all authors, the corresponding author states that there is no conflict of interest to report.

## Acknowledgements

The authors would like to thank Al Green from the Materials Department for his assistance in sourcing supplies essential to the construction of the models. His support and resourcefulness greatly contributed to the successful completion of this project.

**Appendices**

Appendix A. Ear Simulation Model Build Guide

Appendix B. Mirror Laryngoscopy Trainer Build Guide

Appendix C. Nasal and Laryngeal Endoscopy Model Build Guide

Appendix D. Cortical Mastoidectomy Drilling Model Build Guide

Appendix E. Cricothyrotomy Simulation Model Build Guide

Appendix F. Pre- and Post-Workshop Evaluation Surveys

Appendix G. Facilitator Guide

Appendix H. Learner Task Sheets and Checklists

Appendix I. Materials and Cost Summary Tables

