## Appendix I for "A Comprehensive, Low-Cost Multistation ENT Simulation Curriculum for Medical Students: Five Reproducible Task Trainers for Foundational Otolaryngology Skills"

### **Appendix I. Materials and Cost Summary Tables**

#### **Purpose**

To provide a clear, consolidated overview of all materials, quantities, and estimated costs required to construct each of the five low-cost ENT simulation models included in the curriculum. This supports reproducibility, scalability, and ease of implementation for other educators.

### **TABLE 1. Ear Simulation Model — Materials & Costs**

| **Item** | **Quantity per Model** | **Approx. Cost** |
| --- | --- | --- |
| 10 mL syringe (barrel only) | 1 | $0.10 |
| Powder-free glove (finger portion) | 1 section | $0.05 |
| Kidney basin or small cup | 1 | $0.50 |
| Cardboard/foam backing | 1 piece | $0.05 |
| Silicone, peanut butter, or soft compound | Small amount | $0.20 |
| Tape or hot glue | As needed | $0.05 |
| Optional: printed tympanic membrane image | 1 | Free |
| Optional: small beads/seeds (foreign bodies) | 2–3 | <$0.05 |

**Total Estimated Cost:** **~$2.50 per model**

### **TABLE 2. Mirror Laryngoscopy Model — Materials & Costs**

| **Item** | **Quantity per Model** | **Approx. Cost** |
| --- | --- | --- |
| Paper or plastic cup | 1 | $0.10 |
| Cardboard or foam sheet | 1 small piece | $0.05 |
| Printed laryngeal images | 4–6 | Free |
| Glue or tape | As needed | $0.05 |
| Tongue depressor (optional internal support) | 1 | $0.05 |
| Elastic band (optional stabilization) | 1 | <$0.05 |

**Total Estimated Cost:** **~$1.30 per model**

### **TABLE 3. Nasal & Laryngeal Endoscopy (Bell Pepper) Model — Materials & Costs**

| **Item** | **Quantity per Model** | **Approx. Cost** |
| --- | --- | --- |
| Bell pepper | 1 | $0.70 |
| Kidney basin / small bowl | 1 | $0.50 |
| Clear plastic tubing | 1 piece (~1–1.5 ft) | $2.00 |
| Tape | As needed | $0.10 |
| Skewer/dowel (support) | 1 | $0.05 |
| Printed vocal cord image | 1 | Free |
| Small seeds/beads (FB retrieval) | 3–4 | $0.05 |

**Total Estimated Cost:** **~$5.70 per model**

### **TABLE 4. Mastoidectomy Drilling Model — Materials & Costs**

| **Item** | **Quantity per Model** | **Approx. Cost** |
| --- | --- | --- |
| Styrofoam disk (10–12 cm) | 1 | $1.00 |
| Wooden craft ring | 1 | $1.00 |
| Pipe cleaners (critical structures) | 2–3 | $0.30 |
| Glue (wood/hot glue) | As needed | $0.10 |
| Marker | 1 | $0.05 |
| Wooden or plastic base | 1 | $1.00 |

**Total Estimated Material Cost:** **~$3.45**
 **Total Estimated Cost Including Drill Access:** **~$27.00 per model**
 *(Drill & burrs provided by institution; not included in per-model cost.)*

### **TABLE 5. Cricothyrotomy Model — Materials & Costs**

| **Item** | **Quantity per Model** | **Approx. Cost** |
| --- | --- | --- |
| Plastic water bottle | 1 | $0.50 |
| Dry rice/lentils (filler) | ½ cup | $0.20 |
| Household sponge (soft tissue layer) | 1 | $0.30 |
| Disposable glove (Deep fascia layer)  Tattoo Skin (Skin layer) | 1  1 | $0.10  $1.50 |
| Tape (for securing layers) | As needed | $0.10 |
| Plastic tubing (airway) | 1 | $0.50 |

**Total Estimated Cost:** **<$10.00 per model**

### **Global Materials Summary (For Bulk Purchasing)**

| **Model** | **Estimated Cost per Unit** | **Primary Supplies** |
| --- | --- | --- |
| Ear Simulation Model | ~$2.50 | Syringe, glove, peanut butter/silicone |
| Mirror Laryngoscopy Model | ~$1.30 | Cup, printed laryngeal images |
| Bell Pepper Endoscopy Model | ~$5.70 | Bell pepper, tubing |
| Mastoidectomy Drilling Model | ~$27.00 | Styrofoam, wood ring, drill access |
| Cricothyrotomy Model | <$10.00 | Bottle, sponge, glove, rice |
