## Appendix H for "A Comprehensive, Low-Cost Multistation ENT Simulation Curriculum for Medical Students: Five Reproducible Task Trainers for Foundational Otolaryngology Skills"

### **Appendix H. Learner Task Sheets and Skill Checklists**

#### **Purpose**

To provide learners with clear, step-by-step task sheets and procedural checklists for each simulation station. These sheets help learners stay focused, self-direct their practice, and ensure consistency across rotations.

### **Station 1 — Ear Simulation Model: Learner Task Sheet**

##### **Learning Objectives**

- Perform basic otoscopy
- Identify tympanic membrane (TM) landmarks
- Remove cerumen safely
- Retrieve foreign bodies
- Practice TM patching (optional)

#### **Task Checklist**

**Otoscopy**

- ☐ Brace examining hand on model
- ☐ Gently retract “pinna” (model stabilization)
- ☐ Insert otoscope speculum under direct visualization
- ☐ Maintain centered view throughout

**Landmark Identification**

- ☐ Pars tensa
- ☐ Pars flaccida
- ☐ Cone of light
- ☐ Umbo
- ☐ Handle of malleus (printed)

**Cerumen Removal**

- ☐ Use curette with controlled, gentle strokes
- ☐ Maintain visualization while removing cerumen
- ☐ Avoid scraping the “canal” walls

**Foreign Body Retrieval**

- ☐ Identify object
- ☐ Use alligator forceps to grasp
- ☐ Withdraw under visualization

**Tympanic Membrane Repair (optional)**

- ☐ Identify simulated perforation
- ☐ Apply patch with precision

### **📄 Station 2 — Mirror Laryngoscopy Model: Learner Task Sheet**

##### **Learning Objectives**

- Warm and position laryngeal mirror
- Align light source and line of sight
- Depress “tongue” appropriately
- Identify laryngeal structures
- Compare normal and pathological views

#### **Task Checklist**

**Preparation**

- ☐ Warm mirror (no flame)
- ☐ Verify adequate light source
- ☐ Position model at eye level

**Technique**

- ☐ Hold mirror like a pencil
- ☐ Depress “tongue” gently
- ☐ Insert mirror at 45–70° angle
- ☐ Avoid touching posterior “pharynx”

**Anatomy Identification**

- ☐ Epiglottis
- ☐ Arytenoids
- ☐ True vocal cords
- ☐ False cords
- ☐ Interarytenoid area

**Pathology Recognition**

- ☐ Identify pathology images provided
- ☐ Verbalize clinical significance

### **Station 3 — Nasal & Laryngeal Endoscopy Model: Learner Task Sheet**

##### **Learning Objectives**

- Perform rigid nasal endoscopy
- Perform flexible nasolaryngoscopy
- Maintain centered visualization
- Retrieve foreign bodies
- Navigate into simulated “larynx”

#### **Task Checklist**

**Rigid Endoscopy**

- ☐ Grip scope with two-hand stability
- ☐ Insert through “nares” gently
- ☐ Identify nasal septum
- ☐ Locate inferior turbinate
- ☐ Locate posterior choana

**Flexible Endoscopy**

- ☐ Advance scope smoothly
- ☐ Use wrist techniques for steering
- ☐ Transition into tubing (nasopharynx → larynx)

**Foreign Body Retrieval**

- ☐ Locate bead/seed
- ☐ Use forceps with steady control
- ☐ Remove without losing visualization

**Orientation Skills**

- ☐ Demonstrate scope rotation
- ☐ Center the field of view
- ☐ Maintain awareness of model's natural variation

### **Station 4 — Mastoidectomy Drilling Model: Learner Task Sheet**

##### **Learning Objectives**

- Safely handle a rotary drill
- Use correct burr movements
- Identify McEwen’s triangle
- Avoid critical structures (sigmoid sinus, tegmen)

#### **Task Checklist**

**Drill Handling**

- ☐ Secure model
- ☐ Hold drill with stable fulcrum
- ☐ Use sweeping, continuous motions
- ☐ Avoid static contact with foam

**Anatomical Orientation**

- ☐ Identify McEwen’s triangle
- ☐ Recognize simulated sigmoid sinus
- ☐ Recognize tegmen

**Safe Drilling**

- ☐ Stay within cortical boundaries
- ☐ Monitor depth (avoid colored pipe cleaners)
- ☐ Verbalize orientation during drilling

### **Station 5 — Cricothyrotomy Model: Learner Task Sheet**

##### **Learning Objectives**

- Identify thyroid cartilage, cricothyroid membrane, and tracheal rings
- Perform horizontal membrane incision
- Dilate opening
- Insert airway catheter/tube

#### **Task Checklist**

**Anatomy Identification**

- ☐ Palpate thyroid cartilage
- ☐ Locate cricothyroid membrane
- ☐ Identify first tracheal ring

**Procedure Steps**

- ☐ Stabilize “skin” with nondominant hand
- ☐ Make horizontal incision
- ☐ Feel for “pop” entering airway
- ☐ Insert hemostat to dilate
- ☐ Insert catheter/tube into lumen

**Safety and Technique**

- ☐ Maintain midline orientation
- ☐ Keep incision controlled
- ☐ Discuss real-world confirmation techniques
