## Appendix G for "A Comprehensive, Low-Cost Multistation ENT Simulation Curriculum for Medical Students: Five Reproducible Task Trainers for Foundational Otolaryngology Skills"

### **Appendix G. Facilitator Guide for ENT Simulation Workshop**

#### **Purpose**

To provide facilitators with step-by-step guidance for delivering the multistation ENT simulation curriculum, including teaching points, timing, setup instructions, learner coaching strategies, and safety considerations. This guide ensures consistent delivery of the curriculum across instructors and sites.

### **FACILITATOR OVERVIEW**

##### **Workshop Duration:**

**2 hours** total

- Orientation: 10 minutes
- Five rotating stations: 75–90 minutes
- Group debrief: 10 minutes

##### **Learners:**

Third-year medical students completing their otolaryngology rotation.

##### **Group Size:**

3–4 learners per station, rotating every 15–20 minutes.

##### **Facilitator Responsibilities:**

- Demonstrate proper technique at each station
- Provide real-time feedback during learner practice
- Reinforce anatomical orientation and safe instrument handling
- Create an encouraging, low-pressure learning environment
- Guide reflection during the final debrief

### **STATION-BY-STATION FACILITATOR GUIDE**

### **STATION 1: Ear Simulation Model**

##### **Skills Taught**

- Otoscopy technique
- TM landmark identification
- Cerumen removal
- Foreign body extraction
- Basic tympanic membrane patching

##### **Key Teaching Points**

- Brace the examining hand on the patient (or model) for stability.
- Gently straighten the canal (posterior–superior traction).
- Ask learners to identify: umbo, pars tensa/flaccida, cone of light.
- Encourage learners to verbalize findings.
- Demonstrate delicate use of curettes and forceps.

##### **Common Errors to Coach**

- Advancing otoscope too quickly
- Losing visualization while removing cerumen
- Using excessive force with a curette
- Poor hand stabilization

##### **Safety Considerations**

- Remind learners that real patients experience pain with rough technique.
- Practice gentle insertion and minimal pressure.

### **STATION 2: Mirror Laryngoscopy Model**

##### **Skills Taught**

- Mirror warming
- Proper positioning and angle of approach
- Use of tongue depressor
- Light alignment
- Recognition of normal/pathologic laryngeal anatomy

##### **Key Teaching Points**

- Warm the mirror (never with flame for real patients).
- Hold the mirror like a pencil.
- Maintain a clear view of the laryngeal image.
- Align mirror, light, and line of sight.
- Demonstrate gentle, central placement of the mirror.

##### **Common Errors to Coach**

- Fogging due to inadequate warming
- Touching the posterior pharynx
- Poor light alignment
- Excessive pressure on the “tongue”

##### **Pathology Teaching Options**

Use the interchangeable laryngeal images to review:

- Vocal fold edema
- Nodules
- Leukoplakia
- Paralysis

### **STATION 3: Nasal and Laryngeal Endoscopy (Bell Pepper Model)**

##### **Skills Taught**

- Introduction to rigid and flexible endoscopy
- Scope navigation and orientation
- Foreign body retrieval
- Visualization of simulated larynx

##### **Key Teaching Points**

- Demonstrate steady, gentle advancement of the scope.
- Practice rotating the scope to maintain centered visualization.
- Teach “triangle of safety”: stable grip, steady hand, centered field.
- Emphasize spatial orientation when transitioning into tubing (“larynx”).

##### **Common Errors to Coach**

- Rushing forward without maintaining visualization
- Excessive contact with mucosal surfaces
- Losing orientation when entering the tubing
- Using force rather than finesse for FB removal

##### **For Flexible Nasolaryngoscopy**

- Encourage smooth navigation around curves.
- Use small, controlled wrist movements.

### **STATION 4: Cortical Mastoidectomy Drilling Model**

##### **Skills Taught**

- Safe use of a handheld drill
- Burr control and sweeping motions
- Identifying simulated critical structures
- Understanding McEwen’s triangle

##### **Key Teaching Points**

- Always keep the burr **moving**—no static drilling.
- Demonstrate low-pressure, high-control drilling technique.
- Review the boundaries of McEwen’s triangle.
- Identify the simulated sigmoid sinus and tegmen (pipe cleaners).

##### **Common Errors to Coach**

- Digging in one spot
- Drilling too deep too quickly
- Losing orientation while removing “bone”
- Striking simulated critical structures

##### **Safety Considerations**

- Maintain a safe distance between hands and burr.
- Secure model firmly before drilling.

### **STATION 5: Cricothyrotomy Simulation Model**

##### **Skills Taught**

- Identifying thyroid cartilage, cricothyroid membrane, and tracheal rings
- Horizontal membrane incision
- Dilating the airway opening
- Tubing insertion

##### **Key Teaching Points**

- Teach learners to palpate midline structures confidently.
- Emphasize the horizontal incision technique.
- Coach learners to feel for the “pop” of the membrane.
- Reinforce deliberate, controlled dilation and tube placement.

##### **Common Errors to Coach**

- Misidentification of cricothyroid membrane
- Vertical rather than horizontal incision
- Inadequate dilation before tube insertion
- Losing airway alignment

##### **Real-World Considerations**

- Discuss indications, contraindications, pitfalls
- Reinforce calm, step-by-step communication in emergencies

### **FACILITATOR TIMING GUIDE**

| **Component** | **Time** | **Notes** |
| --- | --- | --- |
| Orientation | 10 min | Overview, safety, expectations |
| Station 1 | 15–20 min | Ear procedures |
| Station 2 | 15–20 min | Mirror laryngoscopy |
| Station 3 | 15–20 min | Endoscopy |
| Station 4 | 15–20 min | Mastoidectomy drilling |
| Station 5 | 15–20 min | Cricothyrotomy |
| Group Debrief | 10 min | Review, reflection, key concepts |

### **DEBRIEF GUIDE**

##### **Suggested Debrief Questions**

- “Which station challenged you the most, and why?”
- “What surprised you during the hands-on practice?”
- “Which procedural skills feel more achievable now?”
- “What would you do differently next time with real patients?”

##### **Debrief Themes**

- Growth in procedural confidence
- Importance of spatial orientation
- Awareness of delicate instrument handling
- Value of repetition and low-stakes practice
