## Appendix F for "A Comprehensive, Low-Cost Multistation ENT Simulation Curriculum for Medical Students: Five Reproducible Task Trainers for Foundational Otolaryngology Skills"

### **Appendix F. Pre- and Post-Workshop Evaluation Surveys**

#### **Purpose**

To provide the pre- and post-workshop evaluation instruments used for routine educational program assessment during the ENT simulation curriculum. Surveys measure changes in learner familiarity, procedural confidence, anatomical understanding, and perceived educational value of the simulation stations.

### **PRE-WORKSHOP SURVEY**

**ENT Simulation Workshop: Pre-Workshop Assessment**

All responses are anonymous and used solely for educational program improvement.

##### **Section 1: Baseline Familiarity With ENT Anatomy**

*Please rate your familiarity with the following anatomical areas (1 = Not familiar, 5 = Very familiar).*

1. External ear and tympanic membrane landmarks
    ☐ 1 ☐ 2 ☐ 3 ☐ 4 ☐ 5
2. Nasal cavity and nasopharynx
    ☐ 1 ☐ 2 ☐ 3 ☐ 4 ☐ 5
3. Oral cavity and laryngeal anatomy
    ☐ 1 ☐ 2 ☐ 3 ☐ 4 ☐ 5
4. Anterior neck anatomy (thyroid cartilage, cricothyroid membrane, tracheal rings)
    ☐ 1 ☐ 2 ☐ 3 ☐ 4 ☐ 5
5. Temporal bone and mastoid anatomy (McEwen’s triangle, sigmoid sinus, tegmen)
    ☐ 1 ☐ 2 ☐ 3 ☐ 4 ☐ 5

##### **Section 2: Baseline Procedural Confidence**

*Please rate your confidence performing the following skills (1 = Not confident, 5 = Very confident).*

6. Using delicate ENT instruments (suction, forceps, curettes)
    ☐ 1 ☐ 2 ☐ 3 ☐ 4 ☐ 5
7. Handling a rigid nasal endoscope
    ☐ 1 ☐ 2 ☐ 3 ☐ 4 ☐ 5
8. Handling a flexible nasolaryngoscope
    ☐ 1 ☐ 2 ☐ 3 ☐ 4 ☐ 5
9. Identifying normal otoscopic findings
    ☐ 1 ☐ 2 ☐ 3 ☐ 4 ☐ 5
10. Performing basic mirror laryngoscopy
     ☐ 1 ☐ 2 ☐ 3 ☐ 4 ☐ 5
11. Identifying cricothyroid membrane landmarks
     ☐ 1 ☐ 2 ☐ 3 ☐ 4 ☐ 5
12. Understanding basic mastoidectomy drilling orientation
     ☐ 1 ☐ 2 ☐ 3 ☐ 4 ☐ 5

##### **Section 3: Prior Exposure**

13. Which ENT procedures have you previously observed or performed?
     (Open response)
14. How many ENT procedures have you observed?
     (Open response)

### **POST-WORKSHOP SURVEY**

**ENT Simulation Workshop: Post-Workshop Assessment**

All responses are anonymous and used solely for curriculum improvement.

#### **Section 1: Post-Workshop Procedural Confidence**

*Please rate your confidence with the following ENT skills after completing the workshop (1 = Not confident, 5 = Very confident).*

1. Identifying tympanic membrane landmarks
    ☐ 1 ☐ 2 ☐ 3 ☐ 4 ☐ 5
2. Performing rigid nasal endoscopy
    ☐ 1 ☐ 2 ☐ 3 ☐ 4 ☐ 5
3. Performing flexible nasolaryngoscopy
    ☐ 1 ☐ 2 ☐ 3 ☐ 4 ☐ 5
4. Performing mirror laryngoscopy
    ☐ 1 ☐ 2 ☐ 3 ☐ 4 ☐ 5
5. Identifying cricothyroid membrane landmarks
    ☐ 1 ☐ 2 ☐ 3 ☐ 4 ☐ 5
6. Performing the steps of a cricothyrotomy
    ☐ 1 ☐ 2 ☐ 3 ☐ 4 ☐ 5
7. Understanding mastoidectomy drilling orientation
    ☐ 1 ☐ 2 ☐ 3 ☐ 4 ☐ 5
8. Identifying McEwen’s triangle and mastoid boundaries
    ☐ 1 ☐ 2 ☐ 3 ☐ 4 ☐ 5

#### **Section 2: Educational Value of Each Station**

*Rate the educational value of each model (1 = Not helpful, 5 = Extremely helpful).*

9. Ear simulation model (EAC/TM model)
    ☐ 1 ☐ 2 ☐ 3 ☐ 4 ☐ 5
10. Nose & larynx endoscopy model (bell pepper airway model)
     ☐ 1 ☐ 2 ☐ 3 ☐ 4 ☐ 5
11. Mirror laryngoscopy model
     ☐ 1 ☐ 2 ☐ 3 ☐ 4 ☐ 5
12. Cricothyrotomy model
     ☐ 1 ☐ 2 ☐ 3 ☐ 4 ☐ 5
13. Mastoidectomy drilling model
     ☐ 1 ☐ 2 ☐ 3 ☐ 4 ☐ 5

#### **Section 3: Model Realism & Educational Quality**

14. Anatomical realism of the models
     ☐ 1 ☐ 2 ☐ 3 ☐ 4 ☐ 5
15. Tactile realism / instrument feedback
     ☐ 1 ☐ 2 ☐ 3 ☐ 4 ☐ 5
16. Overall educational value of the curriculum
     ☐ 1 ☐ 2 ☐ 3 ☐ 4 ☐ 5
17. Improvement in confidence from hands-on practice
     ☐ 1 ☐ 2 ☐ 3 ☐ 4 ☐ 5

#### **Section 4: Qualitative Feedback**

18. Which station was the MOST helpful and why?
     (Open response)
19. What improvements would you recommend for any of the models?
     (Open response)
20. Additional comments or feedback:
     (Open response)
