## Appendix E for "A Comprehensive, Low-Cost Multistation ENT Simulation Curriculum for Medical Students: Five Reproducible Task Trainers for Foundational Otolaryngology Skills"

### **Appendix E. Cricothyrotomy Simulation Model Build Guide**

#### **Purpose**

To provide a low-cost, reproducible trainer for teaching fundamental cricothyrotomy skills, including identification of anterior neck landmarks, membrane incision technique, and airway entry. This model allows learners to practice the sequence of emergency front-of-neck access using common materials while providing tactile feedback similar to skin, soft tissue, and tracheal structures.

#### **Materials and Supplies**

| **Item** | **Quantity** | **Approx. Cost** |
| --- | --- | --- |
| Plastic water bottle (with tracheal ridges) | 1 | $0.50 |
| Dry rice or lentils (to fill bottle partially) | ~½ cup | $0.20 |
| Household sponge (to simulate soft tissue) | 1 | $0.30 |
| Disposable glove (cut to deep fascia)  Tattoo Skin (cut to simulate skin) | 1  1 | $0.10  $1.50 |
| Adhesive tape (medical tape or duct tape) | As needed | $0.10 |
| Scalpel or disposable blade | 1 | Institutional supply |
| Plastic tubing or suction catheter | 1 | $0.50 |

**Total estimated cost:** **<$10.00**

#### **Required Instruments for Learners**

- Scalpel
- Kelly clamp or hemostat
- Endotracheal tube, bougie, or catheter (for airway entry practice)
- Gloves

#### **Assembly Instructions**

##### **1. Prepare the Tracheal Framework**

1. Select a plastic water bottle with palpable ridges that can mimic **tracheal rings**.
2. Fill the bottle halfway with dry rice or lentils to provide rigidity and resistance during membrane puncture.
3. Ensure the bottle remains squeezable but firm.

##### **2. Construct the Cricothyroid Membrane Layer**

1. Identify the sloped indentation between two ridges on the bottle—this will represent the **cricothyroid membrane**.
2. Place a small piece of sponge over this area to simulate superficial soft tissue.

##### **3. Create the “Skin”**

1. Cut a section of a disposable glove and stretch it tightly over the sponge.
2. Secure the glove material using medical tape.
3. Ensure adequate tension to replicate the resistance of real skin.
4. Place a sheet of the Tattoo skin to cover the anterior portion of the bottle.

##### **4. Mark Anatomical Landmarks (Optional)**

Use a marker to outline:

- Thyroid cartilage (“Adam’s apple”)
- Cricothyroid membrane
- Tracheal rings

This aids early learners during orientation.

##### **5. Insert the Airway Tube Pathway**

- Ensure a piece of plastic tubing or an open bottle lumen is accessible for learners to insert a catheter or small tube after incision.

#### **Instructions for Learners (Educational Application)**

At this station, learners should complete the full sequence of an emergency cricothyrotomy:

##### **1. Palpation**

- Identify the **thyroid cartilage**, **cricothyroid membrane**, and **tracheal rings**.
- Confirm correct midline alignment.

##### **2. Skin Stabilization**

- Stretch the “skin” taut using the nondominant hand to prevent tissue movement during incision.

##### **3. Horizontal Membrane Incision**

- Make a small horizontal incision through the glove (“skin”) and sponge (“soft tissue”).
- Feel for a subtle “pop” as the scalpel penetrates the simulated cricothyroid membrane.

##### **4. Airway Entry**

- Insert a clamp, dilate gently, and guide plastic tubing or a catheter into the bottle lumen.
- Confirm stable positioning.

##### **5. Discussion of Real-World Adaptations**

- Tube selection
- Ventilation confirmation techniques
- Situational awareness in airway emergencies

#### **Teaching Points for Facilitators**

- Emphasize correct surface anatomy identification—especially distinguishing thyroid cartilage from cricoid cartilage.
- Reinforce the importance of a **horizontal incision** to avoid vascular structures.
- Encourage tactile recognition of membrane puncture (“the pop”).
- Teach learners how to maintain control of the scalpel and stabilize the neck.
- Discuss the indications, contraindications, and potential complications of cricothyrotomy.
- Model calm, step-by-step communication appropriate for high-stakes airway management.

#### **Troubleshooting and Tips**

- **Skin sliding excessively?** Add an extra layer of glove or secure edges more tightly.
- **Membrane too soft?** Use a denser sponge or double-layer.
- **Tube difficult to insert?** Enlarge the incision slightly or widen bottle lumen.
- **Structure too flimsy?** Add additional rice or tape reinforcement.

#### **Time Required for Assembly**

Approximately **5 minutes** per model.

#### **Educational Level**

Appropriate for:

- Medical students learning airway anatomy
- Emergency medicine and anesthesia learners
- ENT trainees beginning airway management instruction
