## Appendix D for "A Comprehensive, Low-Cost Multistation ENT Simulation Curriculum for Medical Students: Five Reproducible Task Trainers for Foundational Otolaryngology Skills"

### **Appendix D. Cortical Mastoidectomy Drilling Model Build Guide**

#### **Purpose**

To provide a practical, reproducible cortical mastoidectomy simulation model that introduces learners to basic drilling mechanics, surface landmark identification, spatial orientation, and early otologic surgical principles. This low-cost model supports deliberate practice in a safe environment and complements supervised operating room exposure. The model design has been previously published as a technical report describing its construction and educational value [9].

#### **Materials and Supplies**

| **Item** | **Quantity** | **Approx. Cost** |
| --- | --- | --- |
| Styrofoam disk (10–12 cm diameter) | 1 | $1.00 |
| Wooden craft ring (to simulate cortical bone boundary) | 1 | $1.00 |
| Pipe cleaners (to simulate sigmoid sinus, tegmen plate) | 2–3 | $0.30 |
| Wood glue or hot glue | As needed | $0.10 |
| Black or colored marker | 1 | $0.05 |
| Small plastic or wooden base | 1 | $1.00 |
| Handheld rotary drill (institution-provided) | 1 | — |
| Various drill burrs (3–5 mm) | Several | — |

**Total cost per unit:** Approximately **$27.00**
 *(Cost includes base components; drill and burrs are institutional equipment.)*

#### **Assembly Instructions**

##### **1. Create the Mastoid Block**

1. Cut or shape a Styrofoam disk to approximate a mastoid cortical surface.
2. Smooth edges to ensure stability and safe handling.

##### **2. Establish the Cortical Boundary**

1. Attach a wooden craft ring to the superior-lateral surface of the Styrofoam using glue.
2. This ring represents the **cortical bone** boundary and provides tactile feedback during drilling.

##### **3. Add Critical Structures**

Construct simulated at-risk structures using pipe cleaners:

- **Sigmoid sinus:**
  - Embed a curved pipe cleaner along the posterolateral quadrant.
  - Position it 3–4 mm beneath the Styrofoam surface.
- **Tegmen plate:**
  - Place a second pipe cleaner superiorly, running anteroposteriorly.
  - Maintain shallow depth to mimic the risk of superior plate violation.

Optional:

- A third pipe cleaner may be placed anteriorly to represent facial recess boundaries.

##### **4. Secure Base Mounting**

1. Glue the assembled Styrofoam model to a small wooden or plastic base for stability.
2. Allow adequate drying time prior to use.

##### **5. Mark Key Orientation Landmarks**

Using a marker:

- Outline **McEwen’s triangle**
- Indicate **superior**, **posterior**, and **anterior** edges
- Label safe vs unsafe drilling zones

These markings assist novice learners in developing intuitive spatial orientation.

#### **Instructions for Learners (Educational Application)**

At this station, learners should practice:

##### **1. Basic Drill Handling**

- Correct hand positioning
- Burr selection (cutting vs diamond burrs)
- Appropriate speed and torque
- Maintaining a stable fulcrum

##### **2. Cortical Bone Removal**

- Begin drilling along the wooden ring boundary
- Use smooth, sweeping strokes
- Maintain continuous awareness of anatomical orientation

##### **3. Identifying Key Structures**

- Observe changing foam texture as “bone” thins
- Recognize pipe cleaners as:
  - **Sigmoid sinus** (posterior)
  - **Tegmen** (superior)
- Practice avoiding these simulated critical structures
- Stop drilling if a colored pipe cleaner becomes visible

##### **4. Understanding McEwen’s Triangle**

- Identify the three bounding lines
- Practice drilling within the safe central zone
- Discuss how this region relates to real cortical mastoidectomy anatomy

#### **Teaching Points for Facilitators**

- Emphasize **low-pressure drilling**, increasing control rather than force.
- Instruct learners to always keep the **burr moving**, avoiding static contact.
- Reinforce the importance of knowing **what lies beneath** each drilling zone.
- Review real surgical correlations:
  - Relationship to sigmoid sinus
  - Tegmen exposure risks
  - Orientation to the mastoid antrum
- Encourage learners to verbalize their orientation continuously.

#### **Troubleshooting and Tips**

- **Drill tearing foam?** Lower speed or increase burr movement.
- **Pipe cleaners exposed prematurely?** Re-explain safe zones and boundaries.
- **Model unstable?** Reinforce base or add sandbag/weight.
- **Learners disoriented?** Revisit McEwen’s triangle markings and start over.

#### **Time Required for Assembly**

Approximately **10–12 minutes** per model.

#### **Educational Level**

Appropriate for:

- Medical students (introductory otologic training)
- Junior ENT residents
- Surgical or EM learners seeking spatial drilling skills
