## Appendix C for "A Comprehensive, Low-Cost Multistation ENT Simulation Curriculum for Medical Students: Five Reproducible Task Trainers for Foundational Otolaryngology Skills"

### **Appendix C. Nasal and Laryngeal Endoscopy Model Build Guide**

#### **Purpose**

To provide a low-cost, reproducible simulation model for teaching rigid nasal endoscopy, flexible nasolaryngoscopy, anatomical landmark identification, and foreign body retrieval. The model recreates the spatial constraints of the nasal cavity and laryngeal inlet using commonly available materials.

#### **Materials and Supplies**

| **Item** | **Quantity** | **Approx. Cost** |
| --- | --- | --- |
| Bell pepper (green or red) | 1 | $0.70 |
| Kidney basin or small plastic bowl | 1 | $0.50 |
| Clear plastic tubing (1–1.5 ft) | 1 | $2.00 |
| Medical tape or duct tape | As needed | $0.10 |
| Spinal needle, skewer, or wooden dowel | 1 | $0.05 |
| Printed image of the vocal cords | 1 | Free |
| Small seeds/beads for foreign bodies | 3–4 | $0.05 |

**Total cost per unit:** Approximately **$5.70**

#### **Instruments for Learners**

- 0° rigid nasal endoscope with light source
- Flexible fiber-optic nasolaryngoscope
- Alligator forceps or ethmoid forceps
- Suction catheter (optional)

#### **Assembly Instructions**

##### **1. Prepare the Pepper**

1. Choose a bell pepper with firm walls and an intact base.
2. Cut off the top portion to expose the internal chamber.
3. Clear seeds and membranes but leave enough structure to provide natural anatomical variability.

##### **2. Create Bilateral Nares**

1. Cut two small oval openings on the anterior surface to represent the left and right nares.
2. Angle these openings slightly upward for realism.

##### **3. Create Nasopharyngeal Openings**

1. On the posterior surface, cut two openings to represent the choanae.
2. These should align with the anterior nares to permit scope passage.

##### **4. Stabilize the Model**

1. Insert a spinal needle, skewer, or dowel into the kidney basin.
2. Mount the pepper upright onto the stabilizing post.
3. Tape the pepper base to prevent rotation during scope maneuvers.

##### **5. Add the “Laryngeal” Pathway**

1. Secure clear tubing to the posterior basin wall using tape.
2. Affix a printed vocal cord image at the distal end of the tubing.
3. Ensure alignment so the flexible scope passes smoothly through the pepper into the tubing.

##### **6. Add Foreign Bodies (Optional)**

- Place small beads or seeds inside the cavity to simulate nasal foreign bodies.

#### **Instructions for Learners (Educational Application)**

Learners should engage in the following activities at this station:

##### **1. Rigid Nasal Endoscopy**

- Insert the 0° scope into the left and right nares.
- Practice gentle advancement while maintaining visualization.
- Identify simulated landmarks such as:
  - Nasal septum
  - Inferior turbinate
  - Middle turbinate (if present)
  - Posterior choana

##### **2. Flexible Nasolaryngoscopy**

- Pass the flexible scope through the nasopharyngeal openings and into the tubing.
- Practice navigating the transition from nasal cavity to pharynx to “larynx.”
- Visualize the printed vocal cords at the tubing terminus.

##### **3. Foreign Body Removal**

- Locate small seeds or beads placed within the pepper.
- Use alligator or ethmoid forceps under direct visualization to remove them.
- Discuss safety considerations for real foreign body situations.

##### **4. Spatial Orientation**

- Practice advancing and withdrawing the scope while keeping the image centered.
- Reinforce hand–eye coordination, instrument steadiness, and scope rotation.

#### **Teaching Points for Facilitators**

- Demonstrate proper gripping technique for both rigid and flexible scopes.
- Reinforce safe instrument entry and avoidance of excessive mucosal contact.
- Coach learners on alignment of scope, light source, and dominant eye.
- Review common nasal pathology and nasal endoscopy indications.
- Discuss how the model’s internal variability mimics real anatomy.

#### **Troubleshooting and Tips**

- **Scope gets stuck?** Enlarge the nasopharyngeal openings slightly.
- **Pepper unstable?** Add extra tape or a clay base for anchoring.
- **Poor visualization?** Adjust light direction or increase insufflation (rigid).
- **Tubing kinked?** Shorten or reposition tubing for smoother passage.

#### **Time Required for Assembly**

Approximately **8–10 minutes** per model.

#### **Educational Level**

Appropriate for:

- Medical students
- PA students
- Early residents
- Emergency medicine learners practicing airway evaluation
