## Appendix B for "A Comprehensive, Low-Cost Multistation ENT Simulation Curriculum for Medical Students: Five Reproducible Task Trainers for Foundational Otolaryngology Skills"

### **Appendix B. Mirror Laryngoscopy Trainer Build Guide**

#### **Purpose**

To provide a low-cost, reproducible mirror laryngoscopy trainer that teaches learners the foundational skills required for indirect laryngoscopy, including mirror warming, hand positioning, tongue control, light alignment, and identification of normal and pathological laryngeal structures.

#### **Materials and Supplies**

| **Item** | **Quantity** | **Approx. Cost** |
| --- | --- | --- |
| Paper or plastic cup (8–10 oz) | 1 | $0.10 |
| Cardboard or foam sheet | 1 small piece | $0.05 |
| Printed laryngeal images (normal + pathology) | 4–6 | Free |
| Glue or tape | As needed | $0.05 |
| Wooden craft stick or plastic tongue depressor | 1 | $0.05 |
| Elastic band (optional) | 1 | <$0.05 |
| Small LED or penlight | 1 | Institutional supply |

**Total cost per unit:** Approximately **$1.30**

#### **Required Instruments for Learners**

- Laryngeal mirror (e.g., size 3 or 4)
- Light source (headlamp, penlight, or overhead lamp)
- Tongue depressor
- Gloves

#### **Assembly Instructions**

##### **1. Construct the Oral Cavity Funnel**

1. Cut the bottom out of a paper/plastic cup to create a cylindrical open-ended “oral cavity.”
2. Optional: Shape or trim the opening to mimic an oral aperture.

##### **2. Add the Posterior Pharyngeal Wall**

1. Cut a small circular or oval piece of cardboard/foam.
2. Glue a printed laryngeal image (e.g., normal vocal cords) onto the surface.
3. Insert this backing into the posterior end of the cup, securing with glue or tape.
4. Leave room to easily swap alternate pathology images.

##### **3. Create Interchangeable Pathology Stations**

1. Print multiple laryngeal images (nodules, edema, paralysis, leukoplakia, etc.).
2. Cut to size and glue onto additional cardboard backings.
3. These can be quickly inserted and removed during teaching.

##### **4. Add Tongue Simulation (Optional)**

- Attach a tongue depressor inside the cup with tape to mimic obstruction or tongue positioning during a real exam.

##### **5. Stabilize the Model**

- Use an elastic band or tape to secure the cup to the table or to a small base.

#### **Instructions for Learners (Educational Application)**

At this station, learners should practice the complete mirror laryngoscopy workflow, including:

##### **1. Preparation**

- Warm the laryngeal mirror using warm water or friction (not flame).
- Adjust the light source for optimal visualization.

##### **2. Positioning**

- Stabilize the light source.
- Hold the mirror like a pencil, angled downward at 45–70 degrees.
- Use the opposite hand to depress the simulated tongue.

##### **3. Visualization**

- Insert the warmed mirror into the simulated oral cavity.
- Avoid touching the back wall (“gag reflex point”).
- Gently rotate the mirror to visualize the laryngeal image.

##### **4. Pathology Recognition**

- Identify normal anatomy on the baseline image:
  - Epiglottis
  - Arytenoids
  - True and false vocal cords
  - Interarytenoid area
- Rotate in pathology images and have learners:
  - Describe findings
  - Compare normal and abnormal states
  - Practice verbalizing their exam

#### **Teaching Points for Facilitators**

- Emphasize mirror warming technique and avoidance of fogging.
- Reinforce correct hand positioning for both mirror and tongue depressor.
- Teach learners to align the light source with their line of sight.
- Practice safe insertion and withdrawal without touching posterior structures.
- Review common laryngeal pathologies and clinical correlations.

#### **Troubleshooting and Tips**

- **Mirror fogging?** Increase warming time or reduce angle of entry.
- **Learners struggling with view?** Adjust light source direction or height.
- **Mirror hitting the “posterior pharynx”?** Coach on gentler insertion path.
- **Poor visualization?** Increase angle between mirror and light; reposition the model.

#### **Time Required for Assembly**

Approximately **5 minutes** per model.

#### **Educational Level**

Appropriate for:

- Medical students
- PA students
- Emergency medicine or ENT interns in early training
