## Appendix A for "A Comprehensive, Low-Cost Multistation ENT Simulation Curriculum for Medical Students: Five Reproducible Task Trainers for Foundational Otolaryngology Skills"

### **Appendix A. Ear Simulation Model Build Guide**

#### **Purpose**

To provide a low-cost, reproducible ear simulation model that enables learners to practice otoscopy, tympanic membrane identification, cerumen removal, foreign body extraction, and basic tympanic membrane repair techniques in a controlled, hands-on educational environment.

#### **Materials and Supplies**

| **Item** | **Quantity** | **Approx. Cost** |
| --- | --- | --- |
| 10 mL syringe (barrel only) | 1 | $0.10 |
| Powder-free glove (latex or nitrile) | 1 finger segment | $0.05 |
| Kidney basin or plastic cup | 1 | $0.50 |
| Cardboard or foam backing | 1 piece | $0.05 |
| Silicone, peanut butter, or modeling compound | Small amount | $0.20 |
| Clear tape or hot glue | As needed | $0.05 |
| Optional: printed tympanic membrane image | 1 | Free |
| Optional: small beads/seeds for foreign bodies | 2–3 | <$0.05 |

**Total cost per unit:** Approximately **$2.50**

#### **Required Instruments (for learners)**

- Otoscope with disposable specula
- Curettes (loop, angled, plastic or metal)
- Suction catheter (e.g., 5 Fr pediatric)
- Alligator or ethmoid forceps
- Cotton swabs

#### **Assembly Instructions**

##### **1. Create the External Auditory Canal**

1. Remove the plunger from a 10 mL syringe and discard the rubber tip.
2. Trim the barrel to the desired length (typically 2–3 cm).
3. Smooth the edges for safety.

##### **2. Construct the Tympanic Membrane**

1. Cut a finger section from a powder-free glove.
2. Stretch it tightly over one end of the syringe barrel to simulate a tympanic membrane.
3. Secure using tape or a thin layer of hot glue.
4. Optional: affix a small circular printed TM image beneath the glove layer for added realism.

##### **3. Prepare the Middle Ear Backing**

1. Cut a small piece of cardboard or foam to the size of the syringe opening.
2. Apply a thin layer of peanut butter, silicone, or modeling compound behind the membrane to simulate middle ear space.

##### **4. Build the Support Base**

1. Position the syringe barrel inside a kidney basin or plastic cup.
2. Use tape or glue to secure the barrel so that the “TM end” faces upward at a slight angle.
3. Ensure the model is stable enough for repeated instrument use.

##### **5. Add Optional Foreign Bodies**

- Insert small beads, seeds, or rice grains into the canal for practice in foreign body retrieval.

#### **Instructions for Learners (Educational Application)**

At this station, learners should:

1. **Perform otoscopy**
   - Insert the otoscope speculum into the canal
   - Advance carefully while adjusting viewing angles
   - Identify major landmarks:
     - Pars flaccida
     - Pars tensa
     - Umbo
     - Cone of light
2. **Practice cerumen removal**
   - Use curettes, cotton swabs, and suction
   - Maintain visualization throughout
   - Discuss gentle vs aggressive scraping force
3. **Foreign body extraction**
   - Use alligator forceps to remove beads/seeds
   - Practice stabilizing the canal with one hand while manipulating instruments
4. **Tympanic membrane repair simulation**
   Practice placing simulated patches over glove membrane defects
   - Emphasize stabilization and fine-motor control

#### **Teaching Points for Facilitators**

- Emphasize safe speculum insertion and hand bracing.
- Reinforce optimal positioning of the otoscope and learner eye alignment.
- Provide coaching on depth perception in a narrow canal.
- Encourage learners to verbalize what they are visualizing during otoscopy.
- Review common real-life otologic findings and pathologies.

#### **Troubleshooting and Tips**

- **TM wrinkles?** Apply more tension to the glove before securing.
- **Canal too wide?** Use a smaller syringe (5 mL) for narrower pediatric canals.
- **Foreign bodies difficult to retrieve?** Add texture (rice or beads) to simulate cerumen impaction.
- **Model unstable?** Add tape or modeling clay to base for improved anchoring.

#### **Time Required for Assembly**

Approximately **5–7 minutes** per model.

#### **Educational Level**

Ideal for:

- Medical students (preclinical and clinical)
- Physician assistant students
- Early residents
